# Increased Risk of New Onset NAION for Initiators of Semaglutide with Type 2 Diabetes in the U.S. Veterans Health Administration

**DOI:** 10.1101/2025.09.15.25335753

**Authors:** Kent Heberer, Adam Bress, Steven Cogill, Ana I. Maldonado, Sun H. Kim, Shriram Nallamshetty, Ying Q. Chen, Mei-Chung Shih, Julie A. Lynch, Jennifer S. Lee

## Abstract

**Importance:** Glucagon-like peptide-1 receptor agonists (GLP-1RAs) are among the safest and most effective medications for diabetes and weight loss and are currently used by millions of individuals worldwide. While their cardiometabolic benefits are well established, emerging observational reports have raised concerns about a potential association between GLP-1 RA use and new-onset non-arteritic anterior ischemic optic neuropathy (NAION).

**Objective:** To emulate a target trial evaluating the risk of NAION associated with initiation of semaglutide, a GLP-1RA, compared with a sodium-glucose cotransporter-2 inhibitor (SGLT2i) as second-line therapy for type 2 diabetes in a nationwide cohort of U.S. Veterans.

**Design:** Active-comparator, new-user, target trial emulation. Marginal cause-specific hazard ratios (HRs) were estimated using overlap weighting to account for confounding. Data analysis was conducted from July 2025 to September 2025.

**Setting:** The Veterans Health Administration (VHA) nationwide health care system between March 1, 2018 and March 1, 2025.

**Participants:** U.S. Veterans with a diagnosis of type 2 diabetes who were current metformin users and had no prior exposure to GLP-1RAs or SGLT2is.

**Exposure:** Initiation of semaglutide or any SGLT2i.

**Main Outcome and Measure:** Incident NAION, identified using ICD-10 and SNOMED diagnosis codes.

**Results:** A total of 102,361 Veterans met inclusion criteria, including 11,478 new initiators of semaglutide and 90,883 new initiators of an SGLT2i. Baseline characteristics were well balanced between treatment groups after overlap weighting (mean [SD] age, 60.1 [11.7] years; BMI, 37.8 [6.7] kg/m^2^; hemoglobin A1c, 7.0% [1.4]; 85.5% male; 61.9% non-Hispanic White; 20.7% Black; 8.1% Hispanic). Over a median follow-up of 2.1 years, 153 total incident NAION events occurred. The incidence rate of NAION was 123 per 100,000 person-years among semaglutide initiators and 67 per 100,000 person-years among SGLT2i initiators. Patients who initiated semaglutide had a 2.33-fold higher risk than patients who initiated SGLT2i (HR, 2.33; 95% CI, 1.54–3.54; P<.001).

**Conclusions and Relevance:** In this nationwide cohort of U.S. Veterans with type 2 diabetes, patients who initiated semaglutide had over a 2-fold increased risk of NAION compared to patients who initiated SGLT2i.

## Introduction

Semaglutide, a glucagon-like peptide-1 receptor agonist (GLP-1 RA), is rapidly growing as a treatment option for type 2 diabetes (T2D) and obesity, with an estimated 15 million U.S. adults using the drug.^1,2^ Although semaglutide is highly effective for improving glycemic control, promoting weight loss, and reducing cardiovascular risk, non-arteritic anterior ischemic optic neuropathy (NAION) has emerged as a concerning serious rare adverse event. Evidence linking semaglutide to NAION remains inconsistent; while some observational studies have reported a positive association,^3–6^ others have found no association.^7–10^ Previous investigations have been limited by heterogeneous data sources and study designs, variable data quality, small number of NAION events, and short follow-up. To address these limitations, we emulated a target trial within the U.S. Veterans Health Administration, the U.S.’s large integrated health care system, leveraging pharmacy dispensing data, detailed clinical covariates, and more than seven years of follow-up.

## Methods

### Study Design

We emulated a target trial evaluating the risk of NAION among patients initiating semaglutide, a GLP-1RA, compared with patients initiating a sodium-glucose cotransporter-2 inhibitor (SGLT2i). We used an active-comparator new user design with propensity score weighting in a retrospective nationwide cohort of U.S. Veterans with T2D. The study was conducted under the CSP2012 study protocol approved by the VA Central IRB (VA Central IRB protocol number 17-24).

### Participants and Exposure

We included U.S. Veterans ≥18 years of age with T2D who were on metformin and initiated semaglutide or an SGLT2i between March 1, 2018, and March 1, 2025 (dual initiators were excluded). The index date was the first outpatient pharmacy fill of either drug. Eligibility required ≥1 ICD-10 codes for T2D within two years before index and a metformin fill within 90 days. Exclusions were prior type 1 diabetes or secondary diabetes diagnosis, exposure to other antidiabetic drugs or >30 days of insulin use within 10 years, and prior SNOMED or ICD9/10 diagnosis codes for NAION, giant cell arteritis, or traumatic optic nerve injury within 10 years of the index date. The treatment strategies were initiation of semaglutide or SGLT2i, based on pharmacy dispensing data.

### Follow-up and Outcomes

Follow-up began on the index date and continued until earliest date among incident NAION, a censoring event of death or competing outcome (giant cell arteritis or traumatic optic nerve injury), or end of study follow-up (July 1, 2025). The primary outcome was incident NAION, defined as ≥1 clinical encounter with a SNOMED diagnosis code (14357004) or ICD-10 code (H47.01).

### Baseline Covariates

Baseline covariates were selected a priori for their potential to confound the association between semaglutide and SGLT2i initiation and NAION risk. Covariates were assessed during the two years before the index date using validated algorithms.

### Statistical Analysis

To address measured baseline confounding, we estimated propensity scores for semaglutide initiation using logistic regression with all baseline covariates included in the model^11^ and implemented them using overlap weighting using the PSweight R package.^12^ Baseline covariate balance was assessed before and after weighting using absolute standardized mean differences, with values <0.01 considered indicative of adequate balance. We calculated both cumulative incidence of NAION (number of events divided by persons at risk) and incidence rates (number of events divided by person-time at risk) by treatment group. Adjusted hazard ratios (HRs) and 95% confidence intervals (CIs) for NAION were estimated using Cox proportional hazards models with overlap weighting, implemented with the PSweight package. Missing baseline covariates with <20% missingness and judged to be missing at random were imputed using multiple imputation with chained equations using the mice R Package,^13^ generating 10 imputed datasets. The primary analysis followed the intention-to-treat principle.

## Results

### Study population

Among 814,019 Veterans who initiated semaglutide or an SGLT2i between March 1, 2018, and March 1, 2025, a total of 102,361 met study eligibility criteria (Figure 1). Of these, 11,478 initiated semaglutide and 90,883 initiated an SGLT2i. Most SGLT2i initiators received empagliflozin (n=90,816). Fewer than 100 initiated dapagliflozin and fewer than 10 initiated canagliflozin.

**Figure 1.**
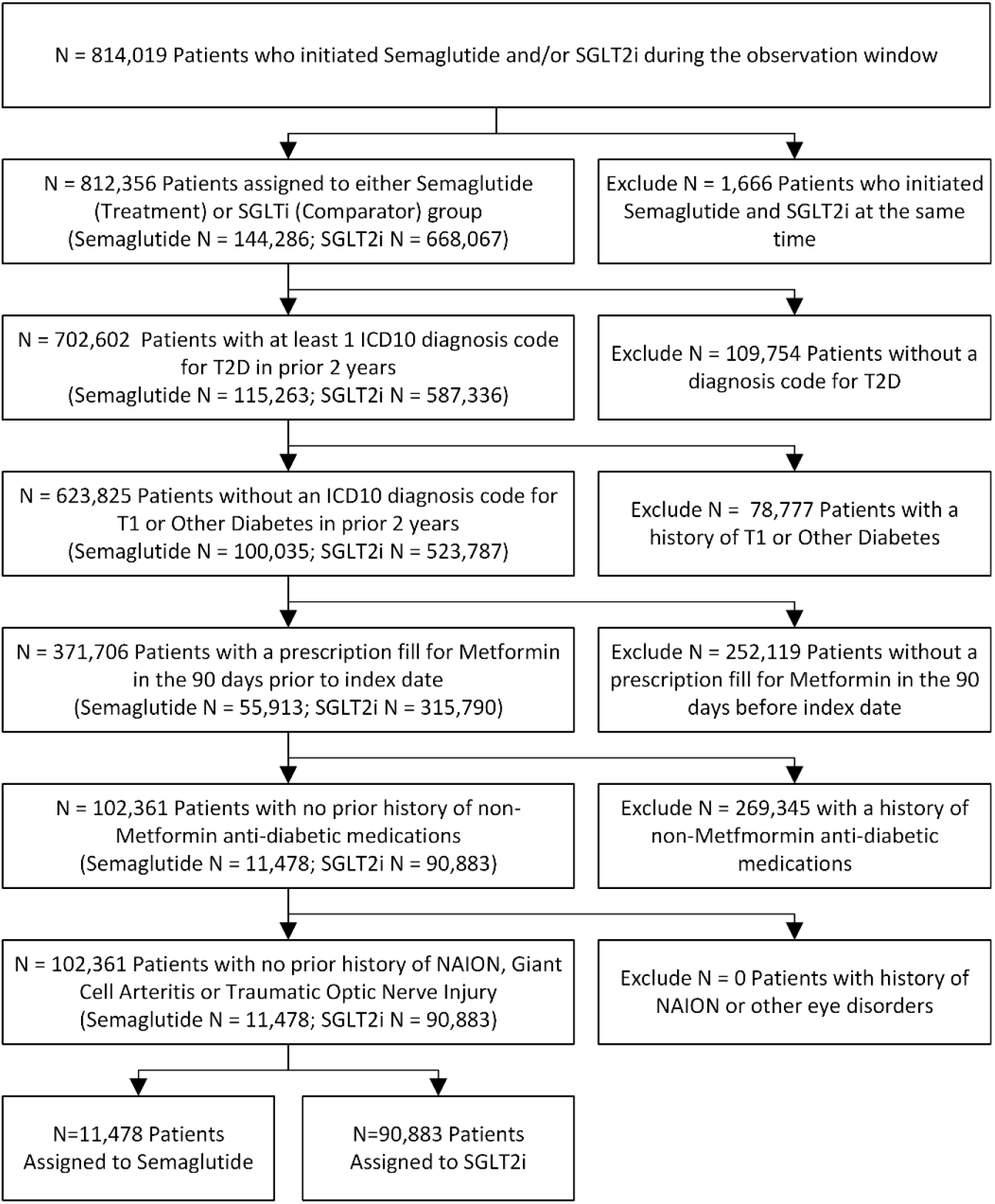
Patient cohort selection flow diagram. Figure depicts the cohort selection logic with inclusion and exclusion criteria, starting with n=814,019 patients who initiated a semaglutide or SGLT2i treatment regimen during the enrollment window and ending with n=102,361 patients in the analytic cohort.

### Baseline characteristics

At treatment initiation, semaglutide users were younger (mean [SD] age, 59.2 [11.7] years vs 64.6 [11.9] years), had higher body mass index (38.8 [7.4] vs 33.6 [6.4]), and had lower hemoglobin A1c (7.0% [1.4] vs 7.3% [1.5]) compared with SGLT2i users. Semaglutide users were also more likely to be female (17.2% vs 6.3%) and Black (21.6% vs 17.8%). After application of overlap weighting, all covariates were well balanced, with standardized mean differences <0.01 (Table 1).

**Table 1.**
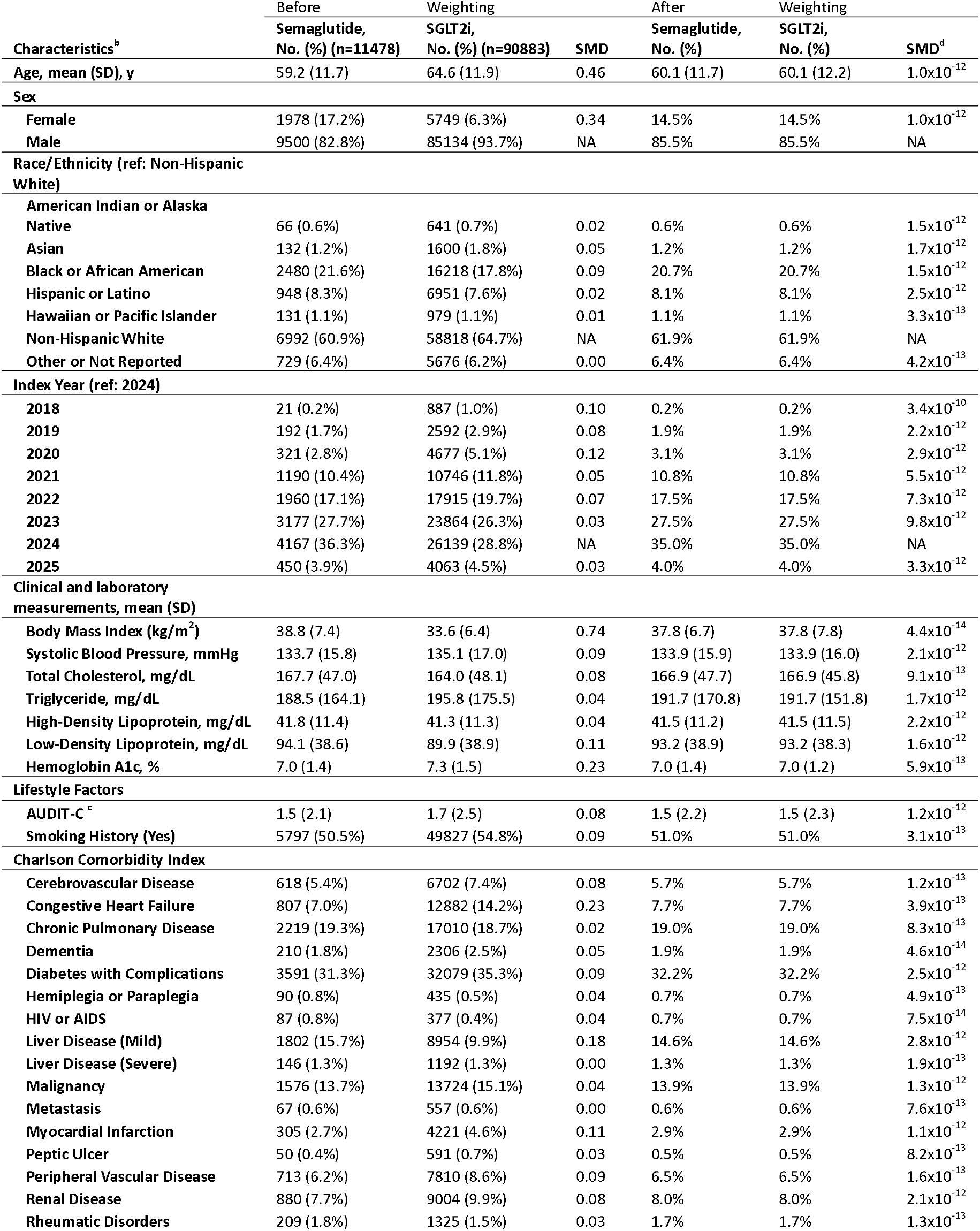
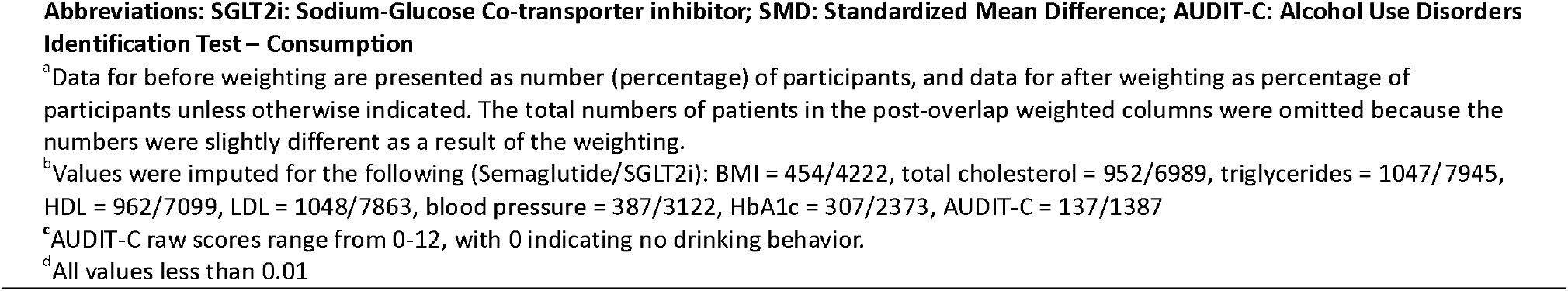
Baseline Covariates Between Initiators of Semaglutide and an SGLT2i Before and After Overlap Weighting^a^

| Characteristics<sup>b</sup> | Before | Weighting | SMD | After | Weighting | SMD<sup>d</sup> |
| --- | --- | --- | --- | --- | --- | --- |
|  | Semaglutide,<br>No. (%) (n=11478) | SGLT2i,<br>No. (%) (n=90883) |  | Semaglutide,<br>No. (%) | SGLT2i,<br>No. (%) |  |
| Age, mean (SD), y | 59.2 (11.7) | 64.6 (11.9) | 0.46 | 60.1 (11.7) | 60.1 (12.2) | 1.0x10<sup>-12</sup> |
| Sex |  |  |  |  |  |  |
| Female | 1978 (17.2%) | 5749 (6.3%) | 0.34 | 14.5% | 14.5% | 1.0x10<sup>-12</sup> |
| Male | 9500 (82.8%) | 85134 (93.7%) | NA | 85.5% | 85.5% | NA |
| Race/Ethnicity (ref: Non-Hispanic White) |  |  |  |  |  |  |
| American Indian or Alaska Native | 66 (0.6%) | 641 (0.7%) | 0.02 | 0.6% | 0.6% | 1.5x10<sup>-12</sup> |
| Asian | 132 (1.2%) | 1600 (1.8%) | 0.05 | 1.2% | 1.2% | 1.7x10<sup>-12</sup> |
| Black or African American | 2480 (21.6%) | 16218 (17.8%) | 0.09 | 20.7% | 20.7% | 1.5x10<sup>-12</sup> |
| Hispanic or Latino | 948 (8.3%) | 6951 (7.6%) | 0.02 | 8.1% | 8.1% | 2.5x10<sup>-12</sup> |
| Hawaiian or Pacific Islander | 131 (1.1%) | 979 (1.1%) | 0.01 | 1.1% | 1.1% | 3.3x10<sup>-13</sup> |
| Non-Hispanic White | 6992 (60.9%) | 58818 (64.7%) | NA | 61.9% | 61.9% | NA |
| Other or Not Reported | 729 (6.4%) | 5676 (6.2%) | 0.00 | 6.4% | 6.4% | 4.2x10<sup>-13</sup> |
| Index Year (ref: 2024) |  |  |  |  |  |  |
| 2018 | 21 (0.2%) | 887 (1.0%) | 0.10 | 0.2% | 0.2% | 3.4x10<sup>-10</sup> |
| 2019 | 192 (1.7%) | 2592 (2.9%) | 0.08 | 1.9% | 1.9% | 2.2x10<sup>-12</sup> |
| 2020 | 321 (2.8%) | 4677 (5.1%) | 0.12 | 3.1% | 3.1% | 2.9x10<sup>-12</sup> |
| 2021 | 1190 (10.4%) | 10746 (11.8%) | 0.05 | 10.8% | 10.8% | 5.5x10<sup>-12</sup> |
| 2022 | 1960 (17.1%) | 17915 (19.7%) | 0.07 | 17.5% | 17.5% | 7.3x10<sup>-12</sup> |
| 2023 | 3177 (27.7%) | 23864 (26.3%) | 0.03 | 27.5% | 27.5% | 9.8x10<sup>-12</sup> |
| 2024 | 4167 (36.3%) | 26139 (28.8%) | NA | 35.0% | 35.0% | NA |
| 2025 | 450 (3.9%) | 4063 (4.5%) | 0.03 | 4.0% | 4.0% | 3.3x10<sup>-12</sup> |
| Clinical and laboratory measurements, mean (SD) |  |  |  |  |  |  |
| Body Mass Index (kg/m<sup>2</sup>) | 38.8 (7.4) | 33.6 (6.4) | 0.74 | 37.8 (6.7) | 37.8 (7.8) | 4.4x10<sup>-14</sup> |
| Systolic Blood Pressure, mmHg | 133.7 (15.8) | 135.1 (17.0) | 0.09 | 133.9 (15.9) | 133.9 (16.0) | 2.1x10<sup>-12</sup> |
| Total Cholesterol, mg/dL | 167.7 (47.0) | 164.0 (48.1) | 0.08 | 166.9 (47.7) | 166.9 (45.8) | 9.1x10<sup>-13</sup> |
| Triglyceride, mg/dL | 188.5 (164.1) | 195.8 (175.5) | 0.04 | 191.7 (170.8) | 191.7 (151.8) | 1.7x10<sup>-12</sup> |
| High-Density Lipoprotein, mg/dL | 41.8 (11.4) | 41.3 (11.3) | 0.04 | 41.5 (11.2) | 41.5 (11.5) | 2.2x10<sup>-12</sup> |
| Low-Density Lipoprotein, mg/dL | 94.1 (38.6) | 89.9 (38.9) | 0.11 | 93.2 (38.9) | 93.2 (38.3) | 1.6x10<sup>-12</sup> |
| Hemoglobin A1c, % | 7.0 (1.4) | 7.3 (1.5) | 0.23 | 7.0 (1.4) | 7.0 (1.2) | 5.9x10<sup>-13</sup> |
| Lifestyle Factors |  |  |  |  |  |  |
| AUDIT-C<sup>c</sup> | 1.5 (2.1) | 1.7 (2.5) | 0.08 | 1.5 (2.2) | 1.5 (2.3) | 1.2x10<sup>-12</sup> |
| Smoking History (Yes) | 5797 (50.5%) | 49827 (54.8%) | 0.09 | 51.0% | 51.0% | 3.1x10<sup>-13</sup> |
| Charlson Comorbidity Index |  |  |  |  |  |  |
| Cerebrovascular Disease | 618 (5.4%) | 6702 (7.4%) | 0.08 | 5.7% | 5.7% | 1.2x10<sup>-13</sup> |
| Congestive Heart Failure | 807 (7.0%) | 12882 (14.2%) | 0.23 | 7.7% | 7.7% | 3.9x10<sup>-13</sup> |
| Chronic Pulmonary Disease | 2219 (19.3%) | 17010 (18.7%) | 0.02 | 19.0% | 19.0% | 8.3x10<sup>-13</sup> |
| Dementia | 210 (1.8%) | 2306 (2.5%) | 0.05 | 1.9% | 1.9% | 4.6x10<sup>-14</sup> |
| Diabetes with Complications | 3591 (31.3%) | 32079 (35.3%) | 0.09 | 32.2% | 32.2% | 2.5x10<sup>-12</sup> |
| Hemiplegia or Paraplegia | 90 (0.8%) | 435 (0.5%) | 0.04 | 0.7% | 0.7% | 4.9x10<sup>-13</sup> |
| HIV or AIDS | 87 (0.8%) | 377 (0.4%) | 0.04 | 0.7% | 0.7% | 7.5x10<sup>-14</sup> |
| Liver Disease (Mild) | 1802 (15.7%) | 8954 (9.9%) | 0.18 | 14.6% | 14.6% | 2.8x10<sup>-12</sup> |
| Liver Disease (Severe) | 146 (1.3%) | 1192 (1.3%) | 0.00 | 1.3% | 1.3% | 1.9x10<sup>-13</sup> |
| Malignancy | 1576 (13.7%) | 13724 (15.1%) | 0.04 | 13.9% | 13.9% | 1.3x10<sup>-12</sup> |
| Metastasis | 67 (0.6%) | 557 (0.6%) | 0.00 | 0.6% | 0.6% | 7.6x10<sup>-13</sup> |
| Myocardial Infarction | 305 (2.7%) | 4221 (4.6%) | 0.11 | 2.9% | 2.9% | 1.1x10<sup>-12</sup> |
| Peptic Ulcer | 50 (0.4%) | 591 (0.7%) | 0.03 | 0.5% | 0.5% | 8.2x10<sup>-13</sup> |
| Peripheral Vascular Disease | 713 (6.2%) | 7810 (8.6%) | 0.09 | 6.5% | 6.5% | 1.6x10<sup>-13</sup> |
| Renal Disease | 880 (7.7%) | 9004 (9.9%) | 0.08 | 8.0% | 8.0% | 2.1x10<sup>-12</sup> |
| Rheumatic Disorders | 209 (1.8%) | 1325 (1.5%) | 0.03 | 1.7% | 1.7% | 1.3x10<sup>-13</sup> |

**Abbreviations: SGLT2i: Sodium-Glucose Co-transporter inhibitor; SMD: Standardized Mean Difference; AUDIT-C: Alcohol Use Disorders Identification Test – Consumption**
<sup>a</sup>Data for before weighting are presented as number (percentage) of participants, and data for after weighting as percentage of participants unless otherwise indicated. The total numbers of patients in the post-overlap weighted columns were omitted because the numbers were slightly different as a result of the weighting.
<sup>b</sup>Values were imputed for the following (Semaglutide/SGLT2i): BMI = 454/4222, total cholesterol = 952/6989, triglycerides = 1047/7945, HDL = 962/7099, LDL = 1048/7863, blood pressure = 387/3122, HbA1c = 307/2373, AUDIT-C = 137/1387
<sup>c</sup>AUDIT-C raw scores range from 0-12, with 0 indicating no drinking behavior.
<sup>d</sup>All values less than 0.01

### Primary outcome

Over 2.1 years median follow-up spanning 239,333 person-years (24,416 for semaglutide, 214,917 for SGLT2i), 30 semaglutide initiators developed NAION (incidence rate, 123 per 100,000 person-years) compared with 143 SGLT2i initiators (67 per 100,000 person-years). Kaplan–Meier curves demonstrated early and persistent separation between groups, with the cumulative incidence of NAION consistently higher among semaglutide initiators (Figure 2). In unweighted analyses, semaglutide initiation was associated with a significantly increased hazard of NAION compared with SGLT2i initiation (HR, 1.84; 95% CI, 1.24–2.73; P=0.002). After overlap weighting, semaglutide initiators had a 2.33-fold higher hazard of NAION (HR, 2.33; 95% CI, 1.53–3.54; P<0.001). The estimated overlap weighted cumulative risk of NAION was 0.29% for semaglutide initiators and 0.12% for SGLT2i initiators.

**Figure 2.**
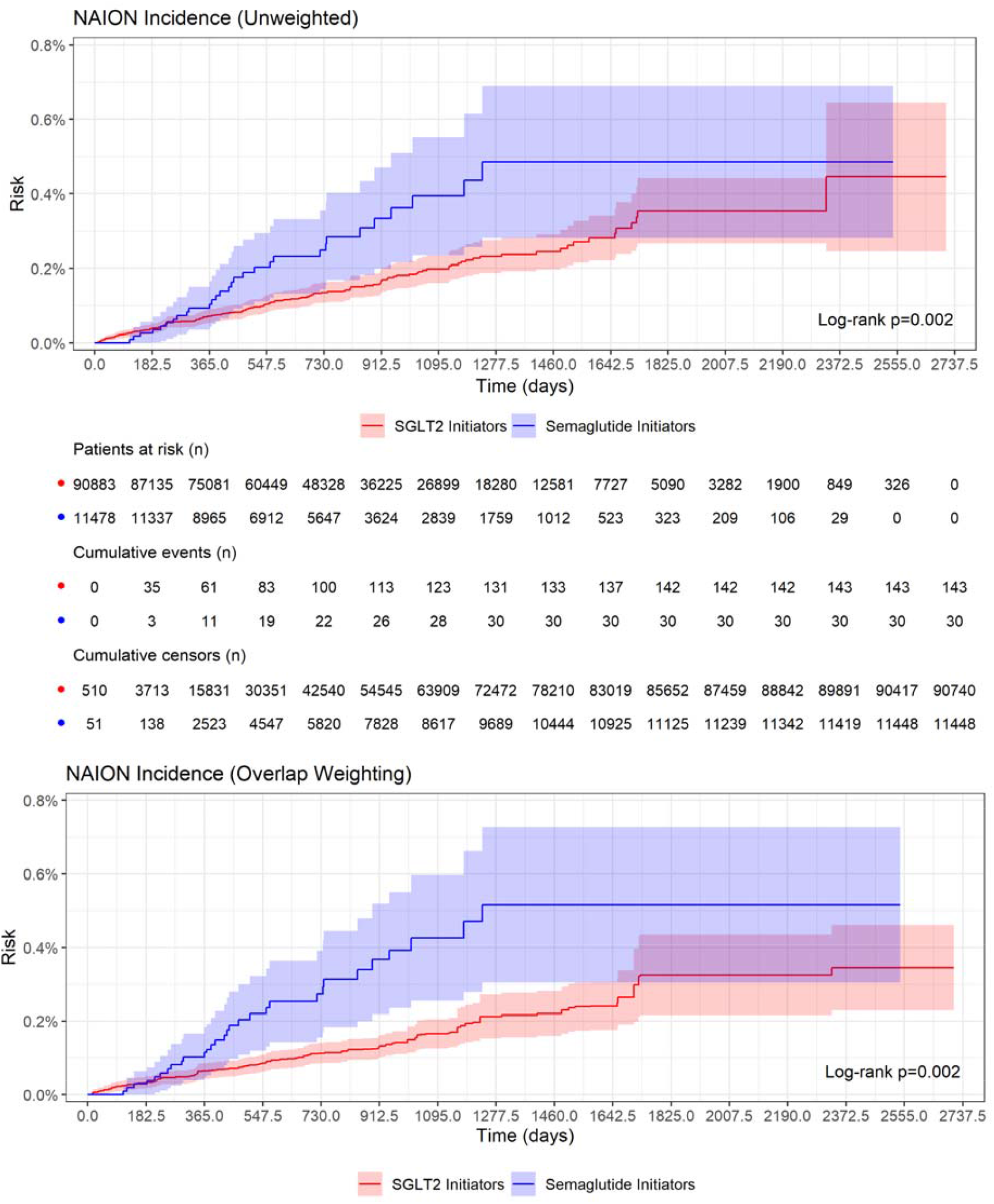
Risk of new-onset NAION in the unweighted cohort (top) and the overlap weighted cohort (bottom). The middle portion shows the number of patients at-risk, number of NAION events, and number of censored events at 6-month intervals for the unweighted cohort. The incidence of NAION is significantly higher in the semaglutide group compared to the SGLT2i group before and after weighting.

## Discussion

In this nationwide target trial emulation of U.S. Veterans with T2D using metformin, initiation of semaglutide was associated with a two-fold higher risk of NAION compared with initiation of an SGLT2 inhibitor after extensive adjustment for clinical covariates using overlap weighting and over a median of 2.1 years of follow-up.

These findings are consistent with the growing body of evidence, including case reports, case series, and population-based studies, suggesting a link between GLP-1 receptor agonist therapy, particularly semaglutide, and NAION. Several large observational analyses have reported similar two-to three-fold risks for semaglutide initiators,^3–6^ whereas others observed attenuated or null associations after extensive covariate adjustment.^7–10^ Importantly, the absolute incidence remains low, translating to roughly one additional case per several thousand treated patients.

The current analysis was designed to address potential reasons for the conflicting findings across prior electronic health record studies. Differences in NAION definitions, comparator choice, use of new-user designs, length of follow-up, referral-based versus population-based sampling, extent of confounder adjustment, and the rarity of NAION may explain prior inconsistencies. This analysis leveraged the U.S.’s largest integrated health system, which offered extended follow-up (up to 7 years) and, by comparison, a relatively large number of incident NAION events.

Methodologically, this study advances prior evidence in several ways. First, medication exposure was defined using outpatient pharmacy dispensing rather than prescription orders or administrative claims. Second, we incorporated detailed clinical covariates, including hemoglobin A1c, body mass index, blood pressure, and lipid levels. Third, our study nearly doubled the semaglutide cohort and accrued six times more at-risk time (24,416 person-years) than the prior OHDSI pooled analysis (6,824 semaglutide initiators [4,429 person-years] and 61,916 empagliflozin initiators [46,496 person-years]).^9^

Limitations include the potential for residual confounding from unmeasured factors. Generalizability may be limited because the Veteran population is older, predominantly male, and has a higher cardiometabolic burden than the general U.S. population.

Additionally, while the positive predictive value of NAION with ≥1 ICD10 code is relatively high using medical records,^14^ miscoding or misdiagnosis could affect overall incidence rates.

The biological mechanism linking GLP-1 RAs to NAION remains unclear. Proposed explanations include nocturnal or postural hypotension, volume depletion from gastrointestinal side effects, rapid glycemic improvement with transient microvascular dysregulation, and impaired vascular autoregulation at the optic nerve head.^15–17^

In conclusion, among Veterans with T2D, initiation of semaglutide was associated with a more than two-fold higher risk of NAION compared with initiation of SGLT2 inhibitor.

Although absolute risk was low, these findings support that semaglutide increases the incidence of NAION and that the consideration of semaglutide may warrant medical counseling about this vision losing event.

## Data Availability

Patient-level data are currently accessible to all VA researchers with appropriate IRB approvals.

## Acknowledgements

This work was supported by using resources and facilities of the Department of Veterans Affairs (VA) Informatics and Computing Infrastructure (VINCI), including manuscript preparation by Kathryn Pridgen, which is funded under the research priority to Put VA Data to Work for Veterans (VA ORD 24-D4V-02).

We thank Dr. Heather Moss, Professor in the Stanford Departments of Ophthalmology and Neurology & Neurological Sciences for her expert insights.

